# Multi-LLM Disagreement as a Scalable Detector of Human Annotation Errors in Structured Data from Clinical Free-Text

**DOI:** 10.64898/2026.05.04.26352392

**Authors:** Stefan Wittlinger, Josefine Meerjansen, Fabian Wolf, Isabella C. Wiest, Matthias P. Ebert, Fabian Siegel, Sebastian Belle

## Abstract

**Objective:** Structured extraction from clinical free-text depends on human annotators whose labels are susceptible to errors and knowledge-driven mistakes; exhaustive quality control is impractical at scale. We evaluate whether disagreement among multiple locally hosted large language models (LLMs) can prioritize human annotations for targeted review.

**Methods:** Multiple LLMs independently extract the same set of structured variables annotated by a human reviewer. For each annotation, an agreement score counts the LLMs matching the human label. Using four locally hosted LLMs (Gemma 3 27B, DeepSeek-R1 70B, GPT-OSS 120B, Mistral Large 3), we evaluated this approach on 910 German-language colonoscopy reports describing endoscopic mucosal resection, with five structured variables per case (anatomical location, two diameters, resection technique, multiple polyps), yielding 4,550 annotations and a 377-case adjudication sample. A stratified sample oversampling low-agreement strata was adjudicated blinded by an experienced reviewer and analyzed with prevalence-adjusted estimates

**Results:** Human error rates rose as LLM agreement fell, from 0% at scores 3–4 to 76% at score 0. The lowest-agreement stratum was only 6.5% of annotations yet concentrated an estimated 80% of errors. The multi-LLM disagreement score achieved a prevalence-adjusted AUC-ROC of 0.991 (95% CI 0.987–0.994) and AUC-PR of 0.893 (95% CI 0.851–0.929) for error detection.

**Discussion:** Multi-LLM disagreement outperformed single models and provided graded operating points for risk-stratified review.

**Conclusion:** Multi-LLM disagreement provides a scalable quality-control signal for targeted review of the highest-yield cases. Because all models run locally, the framework is GDPR-compliant; its language- and task-agnostic design supports application across clinical domains.

## INTRODUCTION

High-quality structured data is the foundation of clinical research, registry-based studies, and machine learning in medicine. In gastroenterology, colonoscopy reports detail polyp characteristics, size, location, resection technique, and count, information critical for quality benchmarking, surveillance planning, outcomes research, and ML applications. Since this is almost exclusively documented as free-text, large-scale structured extraction is a prerequisite for any systematic analysis.

Structured dataset generation from clinical text conventionally relies on human annotators, a process with two main error sources.[1,2] First, annotation is repetitive and high-volume, making lapses, misreading, or oversight common.[3] Second, annotation is typically delegated to less experienced individuals because experienced reviewers lack time for large-scale labeling, and patient privacy requirements often preclude outsourcing to external workers, further limiting who can do the work.

Quality control is correspondingly difficult: double annotation with conflict resolution mitigates random errors but doubles the workload and depends on the second annotator’s quality, while expert review of all labels is the gold standard but impractical at scale, reviewing thousands of cases is precisely the burden that delegation was meant to avoid.

Large language models (LLMs) offer a potential solution and have already been used to extract medical annotations. Prior work on LLM-based extraction from clinical text has mostly focused on comparing individual model accuracy against human annotations, treating LLMs as potential replacements for human annotators[4–6] including work within the field of gastroenterology.[7] Building on this, several studies proposed to improve the quality of such LLM-generated annotations by selectively routing uncertain cases back to human annotators for correction, reducing review burden while improving overall label quality.[8–10]

This can also be reversed, as one can use LLMs to correct and improve already existing human annotation, and reduce the number of cases an experienced reviewer has to review. This builds on ideas from active learning,[11] LLM-based selection,[12] and Confident Learning. [13] As a practical application, Nahum et al. showed that an ensemble of LLMs can be used to surface likely mislabeled examples in existing benchmarks: cases where the ensemble disagreed with the original human annotations were enriched with genuine labeling errors, and correcting them produced an upward shift in reported model performance.[14] Our work extends this intuition to a clinical domain with several distinguishing features. First, we move from curated benchmark datasets to real-world electronic health records, German-language colonoscopy reports of endoscopic mucosal resection, where documentation style, abbreviations, and clinical ambiguity introduce challenges absent from standardized NLP benchmarks. Second, we employ four LLMs hosted entirely on institutional infrastructure, showing that privacy-compliant local models suffice for disagreement-based error detection without cloud LLMs. Third, rather than simply comparing LLM and human labels, we design a stratified sampling and prevalence-adjusted evaluation framework quantifying the predictive value of graded multi-LLM disagreement, enabling targeted review of the highest-yield cases.

In this work, we propose using disagreement between multiple independent LLMs and a human annotator as a signal for prioritizing human labels for review. As a proof of concept, we demonstrate the approach on 910 colonoscopy reports, in which a medical student annotated five structured variables (4,550 annotations total) and four LLMs independently extracted the same variables. The resulting stratification enables efficient adjudication: the lowest-agreement category, comprising only 6.5% of annotations, concentrates an estimated 80% of all errors.

## METHODS

### Ethical approval

Ethical approval was received from Ethics Committee II, Medical Faculty of Mannheim, Heidelberg University (approval number 2021-694). Pseudonymized data processing in retrospective studies is exempt from obtaining individual patient consent under the applicable regulatory framework.

### Dataset and study design

A total of 1,000 cases from the University Hospital Mannheim involving colonoscopy with at least one endoscopic mucosal resection in an inpatient setting were analyzed, spanning the years 2010 to 2014. The dataset was exhaustive, comprising all eligible cases within this period. Sixty cases were excluded due to incomplete reports, yielding a final dataset of 940 cases. Each case was associated with a unique identifier and a free-text colonoscopy report written in German. Of these, 30 cases were reserved for LLM prompt development, leaving 910 cases for evaluation and statistical analysis. The study design is displayed in Supplementary Figure 1.

### Human Annotation

A medical student not previously exposed to the field of gastroenterology served as the primary annotator, extracting five structured categories from each colonoscopy report: (1) anatomical location of the largest polyp, (2) primary diameter in mm, (3) secondary diameter in mm, (4) resection technique (en-bloc vs. piecemeal/fractionated), and (5) whether multiple polyps were removed (yes/no) (see Supplementary Table 1). The annotator worked from the original free-text reports and received initial training on a pilot set of cases. This setup reflects a realistic scenario in clinical research, where medical students are commonly tasked with chart abstraction. Importantly, the medical student was not aware of the ensuing research, as the data had initially been planned for use in a different project.

### LLM-Based Extraction Pipeline

Four LLMs, Gemma 3 27B,[15] DeepSeek-R1 70B,[16] GPT-OSS 120B,[17] Mistral Large 3 [18] were deployed locally on an institutional GPU server, ensuring patient data remained within hospital infrastructure in compliance with data privacy standards. Gemma 3, Mistral Large 3, and DeepSeek-R1 were deployed in Q4_K_M (4-bit block-wise) quantization, while GPT-OSS 120B was deployed in MXFP4 (4-bit mixed floating-point) quantization. Each model received the same system prompt and few-shot examples and was tasked with extracting the identical five categories from each report.

The extraction pipeline ran in sequential stages (Supplementary Figure 2). Each report was processed with few-shot prompting (Supplementary Table 2), and parsed into structured JSON via regex; malformed responses triggered a second LLM repair call. Extracted data were then normalized (cm to mm), expanded into per-polyp records, and the largest polyp selected for analysis. Ties on diameter 1 were broken deterministically by diameter 2 and, if still tied, by order of mention in the report. Anatomical locations were then standardized. Location terms were matched against a predefined reference list (Supplementary Table 3); unmatched terms were mapped by an additional LLM call. Parts of the Python analysis pipeline were developed with assistance from Anthropic’s Claude Opus/Sonnet 4.6.

### Agreement Scoring and Stratified Sampling

For each report and category, an agreement score (0–4) counted the LLMs whose extracted value matched the human label, where 4 denoted full agreement and 0 no agreement. To efficiently evaluate annotation quality across the agreement spectrum, a stratified sampling design was employed.[19] Up to 100 cases were sampled per score for agreement levels 0–2 (max 20 per category) and up to 50 cases per score for levels 3–4 (max 10 per category), oversampling low-agreement strata to ensure statistical power where errors are most likely. For point-by-point adjudication, Gemma 3 27B output was compared against the human annotation in a blinded, randomized review. A single model was required because each case had to present one internally consistent extraction; combining outputs across models (e.g., majority vote) could mix the location and diameter of different polyps and break blinding. Gemma was selected as the comparator because it achieved the highest accuracy on the prompt development set, providing a conservative estimate of human error rates. Importantly, this choice was specific to the evaluation setup and not inherent to the proposed method. In practical deployment, no single-model output is required: the multi-LLM agreement score alone is sufficient to identify high-risk cases, and reviewers can directly consult the original report.

### Adjudication

An experienced doctoral candidate in medicine, familiar with the specific endoscopy reports from previous publications [20,21], but not involved in the primary annotation, adjudicated all sampled cases. For each case, the adjudicator viewed the original report alongside the human and LLM values, presented as “Annotation A” and “Annotation B” in independently shuffled order with the mapping stored separately. The pair was classified as A correct, B correct, both correct, both wrong, or indeterminate; the A/B mapping was applied only after all cases had been classified. We defined two error operationalizations: a strict definition, in which only unambiguously incorrect annotations counted as errors and indeterminate cases (where the report lacked sufficient information, e.g., unstated location or ambiguous removal method) were excluded; and a broad definition, which additionally counted indeterminate cases as errors, since such annotations would still require manual review.

### Statistical Analysis

Inter-annotator agreement was quantified by percentage agreement, pairwise Cohen’s κ,[22] and group Fleiss’ κ.[23] Error rates were reported with Wilson 95% Cis.[24] The predictive value of multi-LLM disagreement was assessed by prevalence-adjusted ROC and PR curves under two error definitions: strict (human error = LLM correct + both wrong) and broad (additionally including indeterminate). 95% Cis for AUC estimates were obtained by stratified bootstrap with 2,000 resamples.[25] The 2.5th and 97.5th percentiles defined the CI bounds, displayed as shaded bands around the curves and as error bars for the Gemma point estimate. To correct for the enriched error prevalence in the evaluation sample, agreement-level weights were defined as the ratio of full-dataset to sample prevalence, and all sample-based analyses were rescaled accordingly to reflect the natural error distribution.

## RESULTS

### LLM-to-Human Agreement

Overall percentage agreement between each LLM and the human annotations, as shown in Supplementary Figure 3, ranged from 80.1% to 94% across the five extraction categories. Agreement was highest for diameter 2 (93.7–94.4%) and diameter 1 (89.8–91.5%), followed by resection status (88.8–92.1%) and multiple polyps (86.8–91.3%), and lowest for location (80.1–82.3%). Combined across categories, agreement ranged from 87.8% (GPT-OSS) to 90.1% (Gemma), a 1.6-point spread indicating similar extraction performance across models, with comparable distributional statistics (Supplementary Figure 4 for location).

### Distribution of Agreement Scores

Score 4 (all four LLMs matched the human) dominated across all five variables, accounting for 71.7–91.5% of cases; scores 3, 2, 1, and 0 ranged from 1.9–10.0%, 1.4–3.7%, 2.0–2.3%, and 3.2–12.3% respectively. Notably, location exhibited the highest proportion of score-0 cases (12.3%), while diameter 2 had the fewest with 3.2%. The results are displayed in Figure 1.

**Figure 1:**
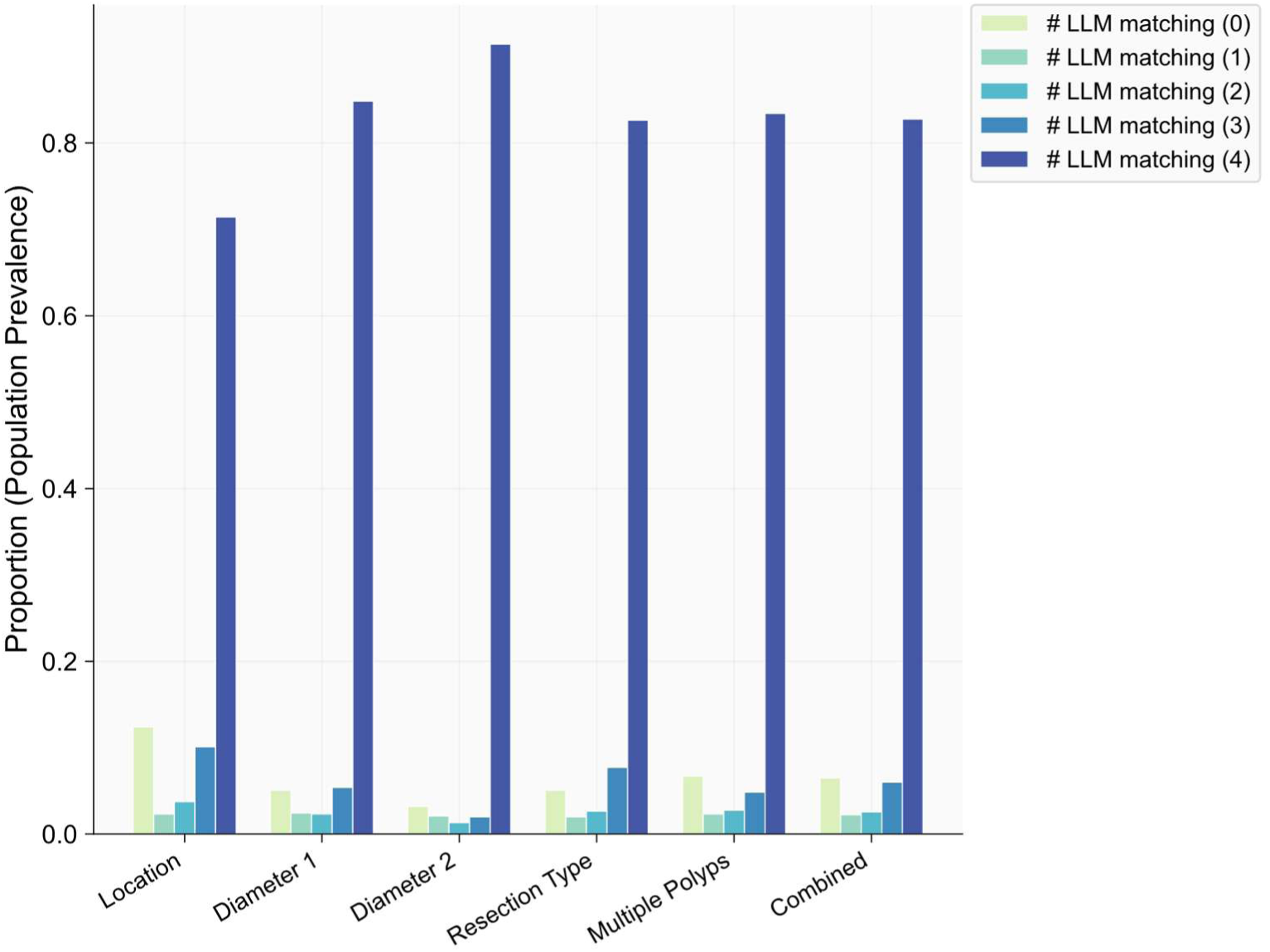
Agreement score prevalence by annotation category. Prevalence of LLM-human agreement scores grouped by annotation category. Each bar represents the proportion of all 910 cases assigned a given agreement score (0–4), where the score denotes the number of LLMs whose extracted label matched the human reference. Alt text: Chart showing, for each of five extraction categories (location, diameter 1, diameter 2, resection type, multiple polyps), the proportion of cases at each agreement score from 0 to 4. Score 4 (all four LLMs match the human) accounts for the large majority of cases in every category at 72–92%, and score 0 ranges from 3% to 12%, with location showing the highest proportion of low-agreement cases.

### Stratified data set

Cases were stratified by number of matching LLMs (Supplementary Figure 5). Groups with 0–2 matches were capped at 100 cases each and groups with 3–4 matches at 50, with up to 20 cases per annotation category (location, diameter 1, diameter 2, resection type, multiple polyps). Cases with IDs below 30 (the prompt-development set) were excluded; combined with limited availability in some categories, the 0–2 match groups reached final totals of 97, 95, and 85 cases respectively, while the 3–4 match groups each reached 50 (10 per category).

### Stratified agreement score

As shown in Figure 2, human error rates rose sharply as LLM agreement fell. Figure 2 a) shows that at a score of 0 (n=97), human error was 76.3%, LLM error 17.5%, indeterminate 6.2%. At score 1 (n=95): human 32.6%, LLM 37.9%, indeterminate 7.4%. At score 2 (n=85): human and LLM both 17.6%, indeterminate 3.5%. At score 3 (n=50): no human errors, LLM error 20.0%. At score 4 (n=50): no errors of either kind. The underlying adjudication results are shown in Figure 2 b) and c). At a score of 0: 74.2% Gemma correct (72/97), 15.5% human correct (15/97), 2.1% both correct, 2.1% both wrong, 6.2% indeterminate. At score 1 (n=95): 41.1% both correct, 18.9% human correct, 13.7% Gemma correct, 18.9% both wrong, 7.4% indeterminate. At score 2 (n=85): 63.5% both correct, 15.3% human correct, 15.3% Gemma correct, 2.4% both wrong, 3.5% indeterminate. At score 3 (n=50): 80.0% both correct, 20.0% human correct. At score 4 (n=50): 100% both correct.

**Figure 2:**
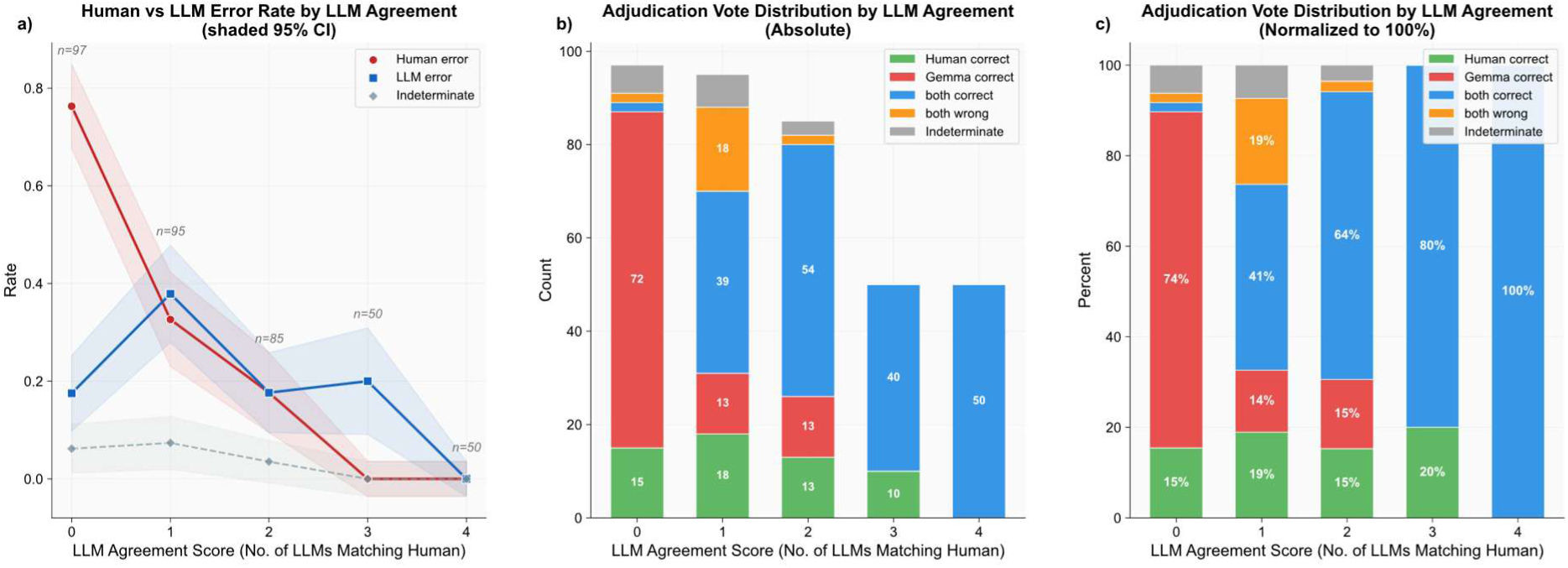
Adjudication results stratified by LLM agreement score. **a)** Human error rate, LLM error rate, and indeterminate rate as a function of the number of LLMs matching the human annotator, with 95% confidence intervals. Sample sizes per group are indicated in gray. **b)** Absolute distribution of secondary reviewer votes across agreement categories (human correct, Gemma correct, both correct, both wrong, indeterminate) for each agreement score. **c)** Normalized distribution of secondary reviewer votes, showing the relative proportion of each category per agreement score. Alt text: Three-panel figure showing how human and LLM error rates change with the LLM agreement score (0–4). Panel a plots human error rate, LLM error rate, and indeterminate rate against agreement score with 95% confidence intervals: human error falls from 76% at score 0 to 0% at scores 3–4. Panels b and c show the absolute and normalized distribution of adjudication outcomes (human correct, Gemma correct, both correct, both wrong, indeterminate) across agreement scores.

### LLM Disagreement as a Predictor of Annotation Errors

Prevalence-adjusted ROC and PR analyses (Figure 3) were performed under two error definitions: strict (LLM correct + both wrong; excluding indeterminate, n=361) and broad (additionally including indeterminate, n=377). Under the strict definition (baseline prevalence 0.063), the four-LLM agreement score achieved weighted AUC-ROC 0.992 (95% CI 0.987–0.995) and AUC-PR 0.888 (95% CI 0.843–0.928). Standalone Gemma reached AUC-ROC 0.945 (95% CI 0.924–0.963) and AUC-PR 0.790 (95% CI 0.742–0.837); as a binary classifier it provides only a single operating point. Under the broad definition (baseline prevalence 0.070), the agreement score achieved AUC-ROC 0.991 (95% CI 0.987–0.994) and AUC-PR 0.893 (95% CI 0.851–0.929); Gemma-only reached AUC-ROC 0.944 (95% CI 0.924–0.961) and AUC-PR 0.801 (95% CI 0.756–0.846).

**Figure 3:**
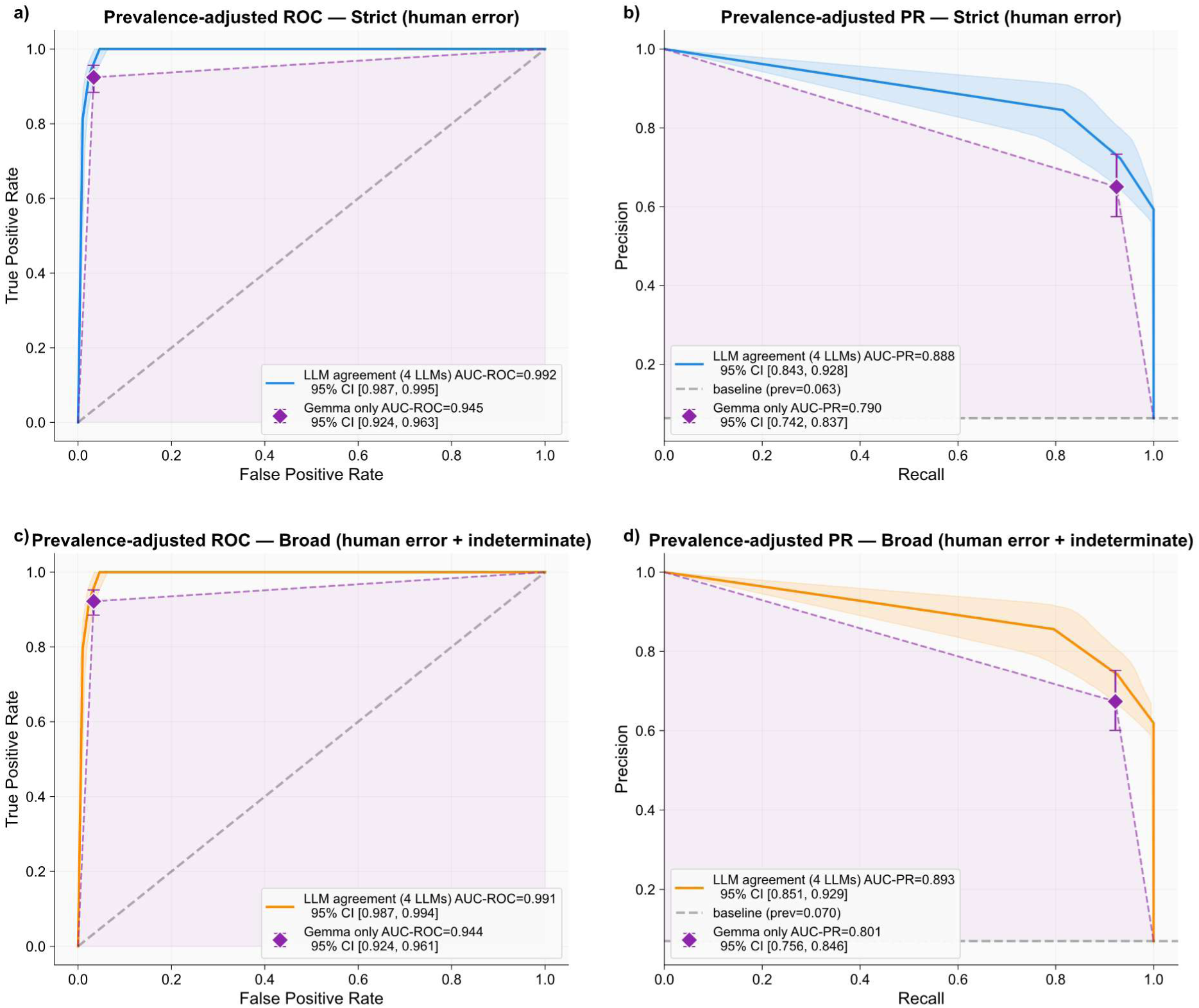
Prevalence-adjusted ROC (left) and PR (right) curves evaluating the predictive value of LLM agreement for detecting human annotation errors under two error definitions. Top row: strict error definition (human error = LLM correct + both wrong, excluding indeterminate cases, n=361). Bottom row: broad error definition (additionally including indeterminate cases as errors, n=377). The solid curves represent the LLM agreement score derived from 4 LLMs, with shaded bands indicating 95% confidence intervals obtained via bootstrapping. The diamond markers denote the single operating point of the binary Gemma output, which, having only a yes/no classification, produces trapezoidal curves (dashed) rather than smooth ones. In the PR plots, the dashed gray baseline corresponds to the prevalence of annotation errors (prev=0.063 for strict, prev=0.07 for broad), representing the precision of a random classifier. Errors are calculated using Wilson confidence intervals. Prev= prevalence. Alt text: Four-panel figure showing receiver operating characteristic curves (left column) and precision-recall curves (right column) under a strict error definition (top row) and a broad error definition (bottom row). Each panel compares the four-LLM agreement score (solid curve with shaded 95% confidence band) against the standalone Gemma classifier (single diamond marker with error bars). Dashed gray lines on the PR plots indicate the baseline prevalence of annotation errors.

### Extrapolated Error

Error rates observed in review were extrapolated to all 4,550 annotations (910 reports × 5 variables), weighted by category prevalence. Figure 4 shows these estimates under both the strict (a–c) and broad (d–f) error definitions. Errors concentrated in the lowest-agreement stratum (panels a, d): at score 0, an estimated 250 errors (strict) and 253 (broad) from a pool of 295 annotations: error rates of 84.6% and 85.6% (panels b, e). This stratum represents 6.5% of all annotations yet contains 82% (strict) and 80% (broad) of all errors. At score 1 (N=101): 36 (strict) and 41 (broad) estimated errors (36.0%, 40.6%). At score 2 (N=116): 20 and 23 errors (17.3%, 20.2%). No errors were observed at scores 3 or 4 (N=273 and N=3,765).

**Figure 4:**
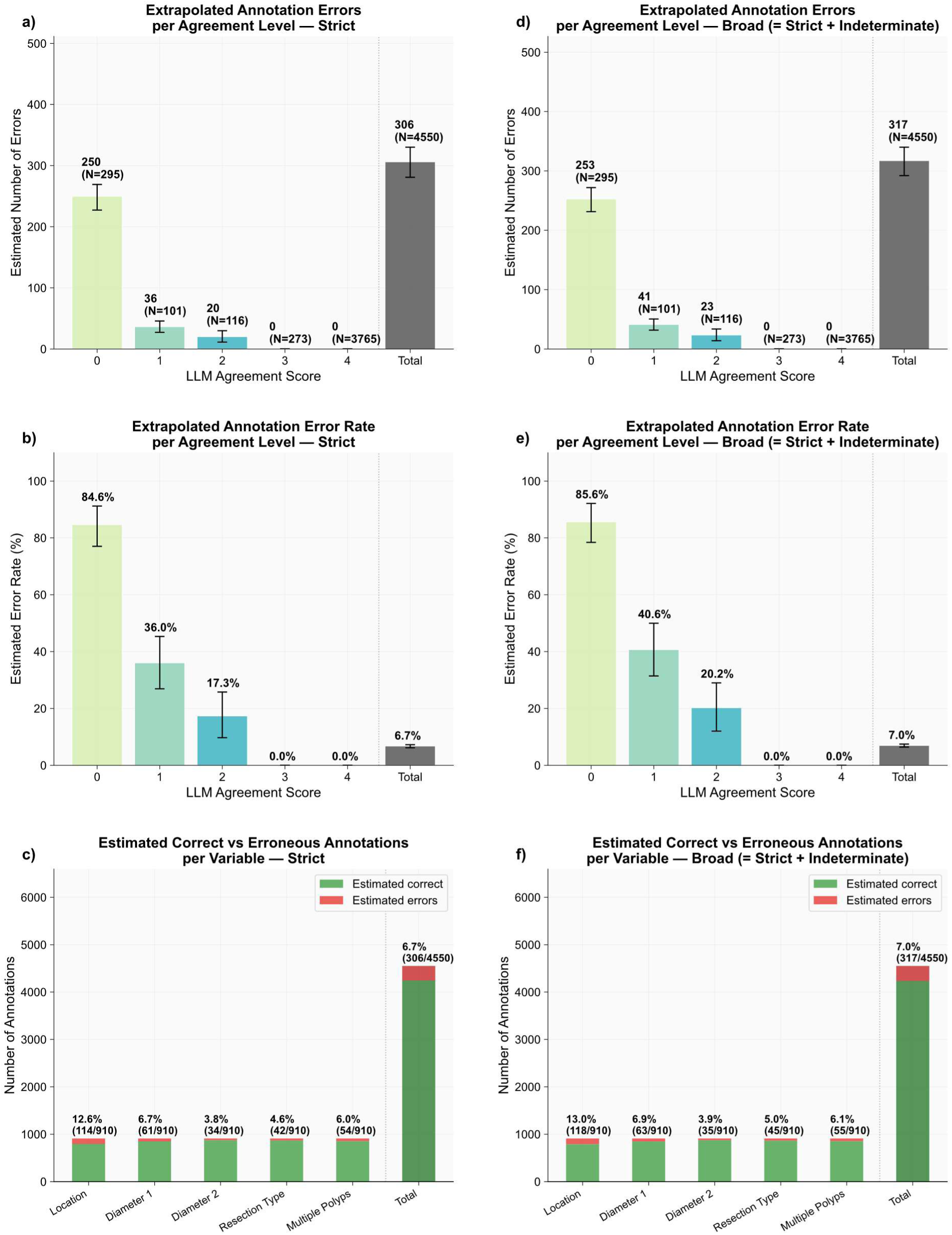
Extrapolated annotation errors across LLM agreement levels and extraction variables. Panels a)–c) show results under the strict error definition, where only annotations that were classified as human error upon secondary review were classified as errors. Panels d)–f) show results under the broad error definition, which additionally counts annotations marked as indeterminate as errors (cases where the reviewer could not definitively determine correctness from the available report text). Error bars represent 95% confidence intervals. a, d) Estimated absolute number of annotation errors per LLM agreement score. The total number of annotations per agreement level is indicated in parentheses (N). b, e) Corresponding estimated error rates (%) per agreement score, i.e. the percentage of errors occurring in a specific agreement class. The overall error rate across all annotations is shown in the rightmost bar. c, f) Distribution of estimated correct (green) and erroneous (red) annotations per extraction variable (each N=910) are shown. Alt text: Six-panel figure projecting error counts and rates from the adjudicated sample to the full dataset of 4,550 annotations. Panels a and d (strict and broad error definitions) show estimated error counts per agreement score, with errors heavily concentrated at score 0 (approximately 250 errors from a pool of 295 annotations). Panels b and e show the corresponding error rates, declining from 85% at score 0 to 0% at scores 3 and 4. Panels c and f disaggregate errors by extraction variable, with location showing the highest error rate (∼13%) and diameter 2 the lowest (∼4%).

Aggregated, the overall estimated error rate was 6.7% (306/4,550) under the strict definition and 7.0% (317/4,550) under the broad. Panels c, f disaggregate by variable: location is highest at 12.6% (114/910; strict) / 13.0% (118/910; broad). The other variables were similar and lower: diameter 1 6.7%/6.9%, diameter 2 3.8%/3.9%, resection type 4.6%/5.0%, multiple polyps 6.0%/6.1% (strict/broad).

### Inter-LLM Agreement

Pairwise inter-LLM agreement (Cohen’s κ) consistently exceeded LLM-to-human agreement across all five categories (Supplementary Figure 6). Location had the lowest overall agreement (inter-LLM κ 0.83–0.89; LLM-to-human 0.82–0.84). Multiple polyps showed the widest gap (inter-LLM 0.88–0.90 vs. LLM-to-human 0.73–0.81), with the lowest human pairwise κ being

0.73 against GPT-OSS. When all five categories were combined into a single composite measure, the human annotator again showed the lowest pairwise κ against each LLM (0.83–0.85), compared to inter-LLM κ values of 0.88–0.92. Fleiss’ κ computed across all four LLMs alone was consistently, though only marginally, higher than Fleiss’ κ computed over all five raters for every extraction category (Supplementary Figure 7).

### Effect of Model Size

Additional experiments with smaller versions of Gemma are shown in Figure 5. Match rates increased substantially with model size, with the most pronounced gains occurring between the 1B and 4B parameter variants. The 1B model performed poorly on structured extraction tasks, particularly for location (17.8%) and diameter 1 (15.9%), while the 4B model achieved markedly higher rates across all categories. Performance differences between the 12B and 27B models were comparatively small. Notably, the 4B model performed similarly to the 12B and 27B variants for diameter 2 and multiple polyps extraction, and only slightly worse for diameter 1, but exhibits substantially lower match rates for resection status (74.5% vs. 87.3%) and location (58.0% vs. 81.0%). Similar results could be obtained for DeepSeek-R1 as shown in Supplementary Figure 8.

**Figure 5:**
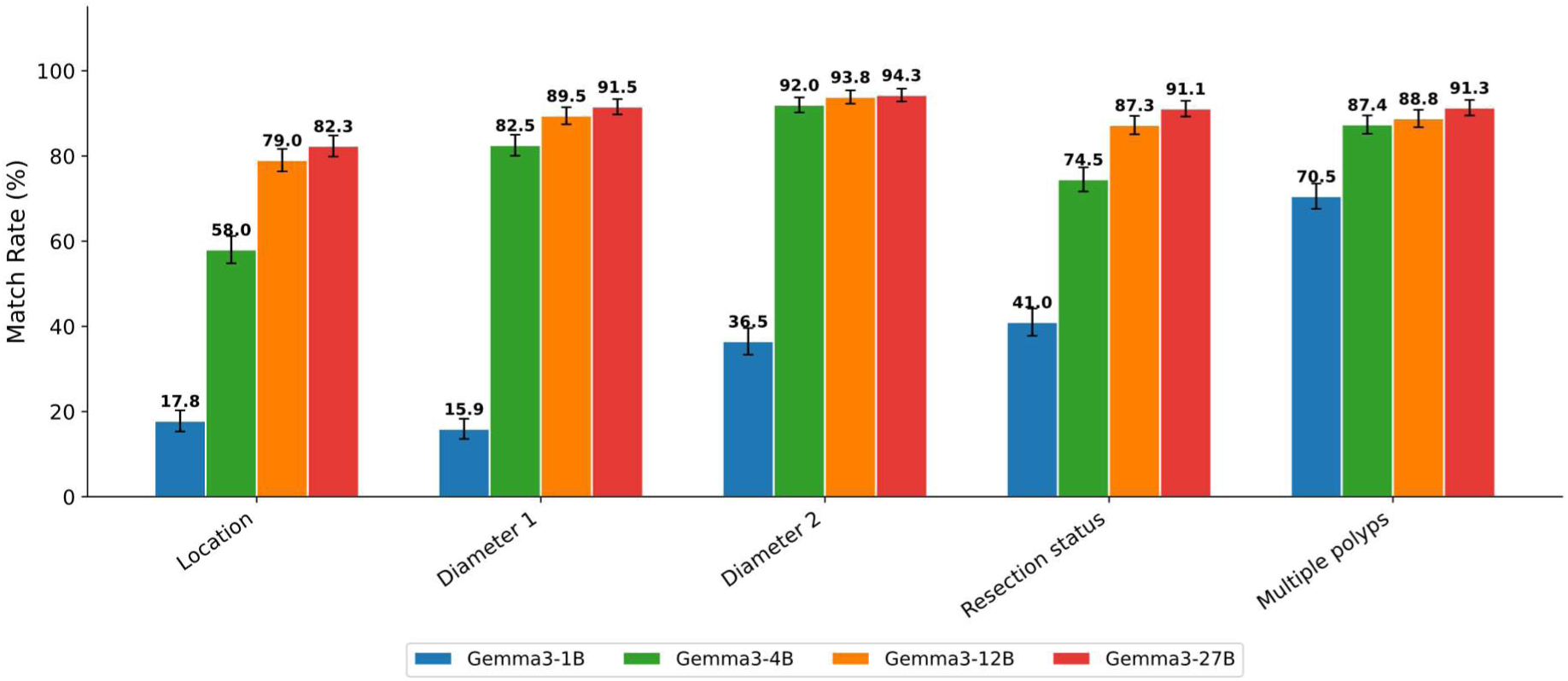
Match rates for different model sizes (Gemma 3). Match rates (%) between LLM-extracted and human-annotated data across Gemma 3 model sizes (1B, 4B, 12B, 27B) for the five extraction targets. Error bars represent 95% confidence intervals. Alt text: Grouped bar chart comparing match rates between LLM-extracted and human-annotated values across four Gemma 3 model sizes (1B, 4B, 12B, 27B parameters) for the five extraction targets (location, diameter 1, diameter 2, resection status, multiple polyps). Match rates rise sharply from the 1B to the 4B variant across all categories, then plateau between the 12B and 27B variants, with location and resection status showing larger gaps between the 4B and 27B models than the other variables.

### Error analysis of Gemma only

Additionally, instead of using stratification, the results for a single LLM were analyzed using only the match/no-match distinction, in this case Gemma 3 27B (Supplementary Figure 9). Among the 179 cases under review where Gemma and the human annotator disagreed, the adjudication sided with the LLM in 54.7% of cases (raw) and with the human annotator in only 31.3%. Both the LLM- and the human-annotator were judged correct in 5.0% of cases, both were wrong in 1.7%, and 7.3% remained indeterminate. After prevalence adjustment, these numbers changed only marginally (59.6% LLM correct, 28.8% human correct, 3.8% both correct, 1.1% both wrong, 6.7% indeterminate). In contrast, prevalence adjustment had a substantial effect on the agreement cases: when Gemma and the human annotator agreed (n=198), the reviewer confirmed both as correct in 88.9% of raw cases (99.4% after adjustment), while both were wrong in 9.6% (0.5% after adjustment).

### Reason for LLM-mismatch

The 56 cases in which Gemma 3 27B produced incorrect classifications while human annotation was correct were subjected to qualitative error analysis. For each case, the model’s output was reviewed alongside the corresponding endoscopy report to identify the underlying reason for misclassification. This analysis yielded four distinct error categories. The corresponding distribution is illustrated in Supplementary Figure 10. The most prevalent error category was incorrect size identification, accounting for 22 cases (39%). The second most common category was re-resection classified as new polyp, observed in 16 cases (29%). Polyp not detected accounted for 10 cases (18%), and wrong location identification was observed in 8 cases (14%). To provide concrete examples of the error categories identified above, four representative examples of Gemma 3 27B misclassifications are presented in Table 1, each corresponding to a different error category. These examples were selected qualitatively to demonstrate the range of failure modes observed and are intended to give the reader an intuitive understanding of how specific report characteristics led to incorrect LLM output.

**Table 1:** Examples where human annotation was correct and Gemma was wrong. The four error types where a distinct LLMs error could be identified (size incorrectly identified, re-resection classified as new polyp, polyp not detected, and wrong location identified) are each illustrated with an example of why Gemma failed to classify correctly.

| Error Type | Real example |
| --- | --- |
| Size incorrectly identified | Three lesions are described with a size range of 10–20 mm; the LLM output 10 mm for the second diameter instead of 0 mm, as it did not recognize that the range denotes a single dimension rather than two distinct diameters. Where no second diameter is explicitly stated, the correct value is 0 mm by definition. |
| Re-resection classified as new polyp | A small residual adenoma margin was removed separately; the LLM counted it as a second independent polyp instead of recognizing it as a fractional re-resection of the same lesion. |
| Polyp not detected | A small polyp bud was explicitly resected with a snare but was not retrieved from the colon; the LLM excluded it because it was not retrieved, failing to recognize that resection and retrieval are distinct, i.e. the polyp had already been removed. |
| Wrong location identified | The scope was advanced to the colon transversum, but the polyp was found at 30 cm from the anus during withdrawal; the LLM assigned 'Colon transversum' (the insertion endpoint) as the location instead of the explicit distance value stated in the text. |

## Discussion

This work proposes multi-LLM disagreement as a general quality-control signal for human annotations in structured clinical extraction, evaluated here on colonoscopy reports. The sheer volume and repetitiveness of chart abstraction tasks make incidental errors, typos, copy-paste mistakes or momentary lapses in attention a common error source. In clinical research, annotation is often delegated to less experienced individuals (e.g., medical students) because experienced clinicians lack time for large-scale labeling, and a first-time annotator may lack the clinical context to interpret ambiguous findings. In an analysis of 10 data sets, an error rate of at least 3.3% was found.[26] A recent systematic review and meta-analysis of 93 medical-record-abstraction studies found a pooled error rate of 6.57%,[27] Our 6.7–7.0% error rate across 4,550 annotations aligns closely with this benchmark.

Traditional quality control (double annotation, random sampling, post-hoc auditing) is labor-intensive and often impractical: double annotation doubles the cost of an already resource-constrained process, and random audits allocate review uniformly rather than toward cases most likely to contain errors. Even with domain experts, inter-rater variability remains substantial regardless of experience level [28,29], indicating human annotation is inherently error-prone. Crowdsourcing, effective in some NLP settings,[30,31] conflicts with medical privacy requirements.

Our approach inverts this paradigm: the LLM agreement score identifies likely-error cases, focusing review on the highest-yield subset and capturing both systematic misinterpretation and sporadic errors that random sampling would miss. The lowest-agreement category, only 6.5% of annotations, contains the majority of estimated errors, a feasible review volume. This shifts quality control from uniform to risk-stratified review, analogous to pre-test-probability-based screening.

Even standalone, Gemma 3 27B achieved solid error detection (AUC-ROC 0.945, AUC-PR 0.790 at low baseline prevalence), showing a single LLM can serve as a useful QC tool. The multi-model agreement score, however, substantially improved discrimination: disagreement from several models is a strong indicator of human error. When all four models agreed with the human label, the annotation was almost always correct, so multi-LLM consensus functions both as an error flag and as a confirmation signal, letting reviewers safely exclude a large fraction of annotations and focus on genuine disagreements. Accordingly, the four-model score consistently outperformed standalone Gemma in weighted AUC-PR under both error definitions.

Multiple LLMs also offer a practical advantage. Standalone Gemma is binary: one operating point; the graded score offers five levels (0–4 LLMs agreeing), yielding multiple operating points along the ROC and PR curves. Users can tune the sensitivity–PPV trade-off to their resources: high-sensitivity thresholds when reviewers are plentiful, stricter thresholds when capacity is limited.

The lower agreement and higher error rates for anatomical location likely reflect how location is documented. Unlike diameter or resection technique, location can be a named segment, a distance from the anal verge, a landmark, or a combination, with German-language reports adding variability through abbreviations and inconsistent naming. Location is also frequently not stated alongside the polyp but must be inferred from preceding sentences, requiring both annotators and LLMs to track context across statements rather than extracting an in-place value.

Besides human error, LLM failures were observed. Qualitative analysis of incorrect Gemma 3 output identified four typical modes. The most common was incorrect size identification (e.g., treating “10–20 mm” as two diameters rather than a range). The second was re-resections misclassified as new polyps, a clinically nuanced distinction. Whether these reflect fundamental LLM limits or insufficient prompting remains open, though the systematic patterns suggest targeted prompt engineering could mitigate them.[32] Reflecting realistic deployment, prompts were developed on 30 cases and applied without iterative refinement against the test set. This underscores anticipatory prompt design, a clinician familiar with the source material is better positioned to anticipate interpretive pitfalls and encode disambiguation into the prompt. Effective LLM-based annotation, and by extension disagreement-based error detection, therefore depends not only on model capability but on prompt quality.

Experiments across model sizes for both Gemma 3 (1B–27B) and DeepSeek-R1 (1.5B–70B) confirmed that agreement rates increase with model size, consistent with established scaling laws.[33,34] Since nearly all cases where top-end models matched were also correctly classified, a lower agreement rate implies a higher false-positive rate. Beyond a model-family-specific threshold, approximately 12B for Gemma 3 and 14B for DeepSeek-R1, further increases in model size yielded only marginal improvements in agreement. For institutions running LLMs locally with limited GPU resources, this suggests that mid-range models might offer a sufficient trade-off between accuracy and computational cost, and that deploying the largest available model is not always necessary.

A key strength is that reports were in German, whereas most prior work on LLM-based clinical extraction used English data.[35,36] Clinical documentation varies across languages in vocabulary, structure, abbreviation patterns, and implicit conventions, yet the strong performance here suggests multi-LLM disagreement as a QC signal is robust to such variation and may generalize beyond English-centric settings. In practice, the framework can be applied to any annotated dataset (see Figure 6); locally hosted inference frameworks not requiring programming expertise can facilitate adoption,[37] making it accessible to clinical researchers. Local deployment keeps patient data within institutional infrastructure, compatible with data protection regulations such as GDPR.[38]

**Figure 6:**
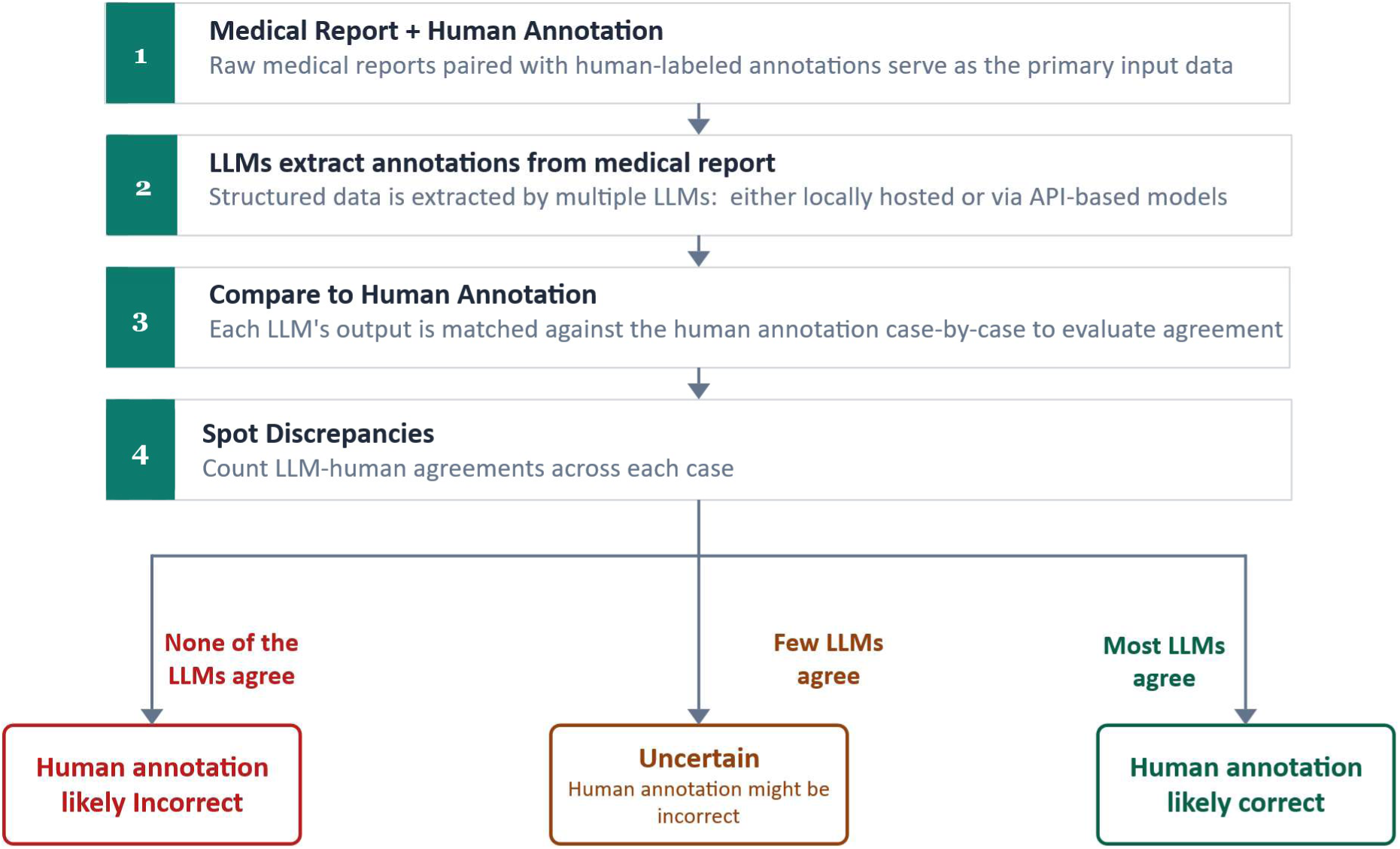
Usage of inference pipeline in a real-world setting. The workflow proceeds in four steps: **(1)** existing annotated records are processed by multiple independent large language models, each of which independently extracts structured information from the source text; **(2)** the resulting LLM outputs are compared against the original human annotations, and discrepancies are flagged; **(3)** the more LLMs disagree with a human annotation, the higher its prioritization for review, under the assumption, supported by our data, that disagreement magnitude correlates with the likelihood of genuine annotation error; **(4)** reviewers concentrate their effort on this high-priority subset rather than re-examining the entire dataset. Importantly, this workflow is not specific to the clinical domain or extraction task studied here; it constitutes a general-purpose quality control mechanism applicable to any setting in which structured annotations are derived from unstructured source material. Local inference frameworks can facilitate adoption by allowing institutions to run this pipeline on their own infrastructure without requiring programming expertise or external data transfer. Alt text: Schematic diagram of a four-step quality-control workflow. Step 1: existing annotated records are processed in parallel by multiple independent large language models, each producing structured extractions. Step 2: LLM outputs are compared against the original human annotations and discrepancies are flagged. Step 3: each annotation receives an agreement score reflecting how many LLMs disagree, with lower agreement indicating higher likelihood of human error. Step 4: reviewers concentrate effort on the high-priority subset rather than the entire dataset.

Several limitations apply. First, this is a single-center study using German colonoscopy reports, limiting generalizability to other settings, languages, or documentation styles; whether the disagreement–error relationship holds across institutions, EHR systems, or report formats requires multi-center validation. Second, these extraction tasks are a specific subset; tasks requiring more complex clinical reasoning or greater linguistic ambiguity may yield different disagreement–error dynamics. Third, the study used a single medical student as primary annotator and a single experienced reviewer for adjudication. The single annotator precludes distinguishing systematic from random errors; the single (blinded, dataset-familiar) adjudicator precludes assessing inter-adjudicator agreement. Fourth, LLM performance depends on prompt design and model selection; we did not systematically explore prompt design, and proprietary cloud-based LLMs were not evaluated because transmitting clinical data externally was incompatible with institutional data protection requirements.

Future work should evaluate this framework across institutions, languages, and clinical domains beyond gastroenterology—radiology, pathology, or other clinical text, since any structured extraction task with multiple independent LLM labels is suited to disagreement-based error detection. Extending to multiple prompts for a single model could further improve detection. The findings also suggest an efficient annotation pathway: cases with full LLM agreement accepted with minimal review, concentrating human effort on disagreements. Evaluating this hybrid strategy and operationalizing the framework through accessible tools would lower adoption barriers.

## CONCLUSION

Disagreement between multiple locally hosted LLMs and a human annotator provides a reliable, scalable signal for detecting annotation errors in structured extraction from colonoscopy reports. The lowest-agreement category (6.5% of annotations) concentrated an estimated 80% of all errors, and the multi-LLM score achieved a prevalence-adjusted AUC-ROC of 0.991 and AUC-PR of 0.893. Stratifying by LLM-to-human disagreement directs secondary review to the highest-yield cases, enabling large high-quality clinical datasets with substantially reduced review effort. Because the framework requires only locally hosted open-weight models and is agnostic to language, domain, and extraction task, it offers a practical, privacy-compliant quality control mechanism for clinical research at scale.

## Data availability

The data are not publicly available because they consist of electronic health records collected at the University Hospital Mannheim. Publicly sharing this data violates the terms of the original ethical approval and could compromise patient privacy. De-identified patient data or other prespecified data will be made available upon approval of a written request and the signing of a data sharing agreement.

## Code availability

The underlying code of this project is available on GitHub (https://github.com/stewitt/llm_clinical_annotation_analysis). Data extraction and statistical analyses were performed using Python (version 3.10.11) with the pandas (version 2.3.2),[39] SciPy (version 1.12.0),[40] NumPy (version 1.26.4),[41] and scikit-learn (version 1.7.2) libraries.[42] LLM inference was performed using the OpenAI Python client library (version 2.16.0).

## Contributions

Stefan Wittlinger (Conceptualization, Methodology, Software, Formal analysis, Investigation, Data curation, Visualization, Writing—original draft, Writing—review & editing), Josefine Meerjansen (Data curation, Investigation), Fabian Wolf (Conceptualization, Methodology, Writing—review & editing), Isabella C. Wiest (Conceptualization, Methodology, Writing—review & editing), Matthias P. Ebert (Resources, Writing—review & editing), Fabian Siegel (Conceptualization, Resources, Writing—review & editing), and Sebastian Belle (Conceptualization, Methodology, Supervision, Writing—review & editing).

## Supplementary Material

**Supplementary Figure 1:**
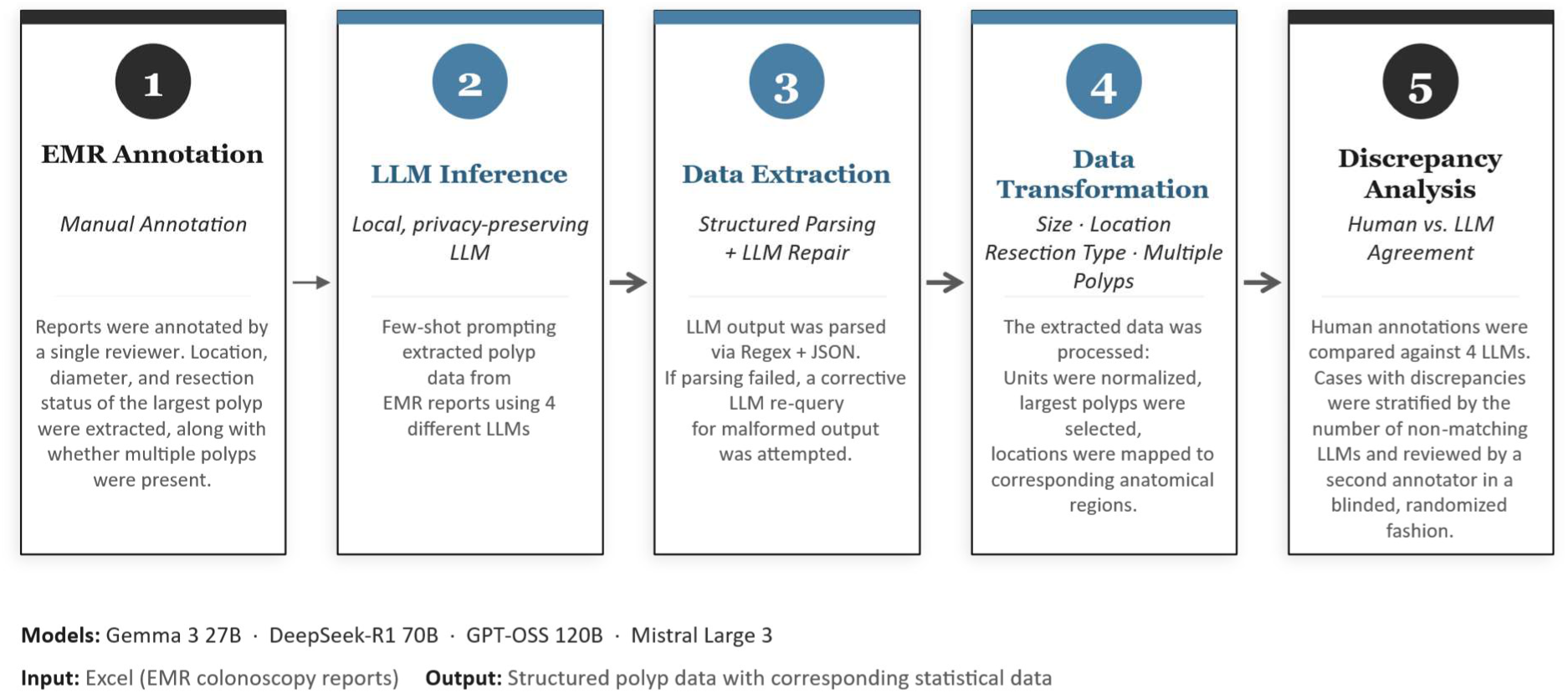
Study design. (1) The dataset was annotated by a single human. Four LLMs were then deployed to flag potential errors in the human annotations of EMR reports. The pipeline comprised (2) few-shot prompted inference to extract polyp characteristics from free-text reports; (3) structured JSON extraction with regex-based parsing and automated LLM re-query as fallback for malformed outputs; (4) data transformation including unit normalization, largest-polyp selection, and anatomical mapping; and (5) discrepancy analysis of potential errors.

**Supplementary Figure 2:**
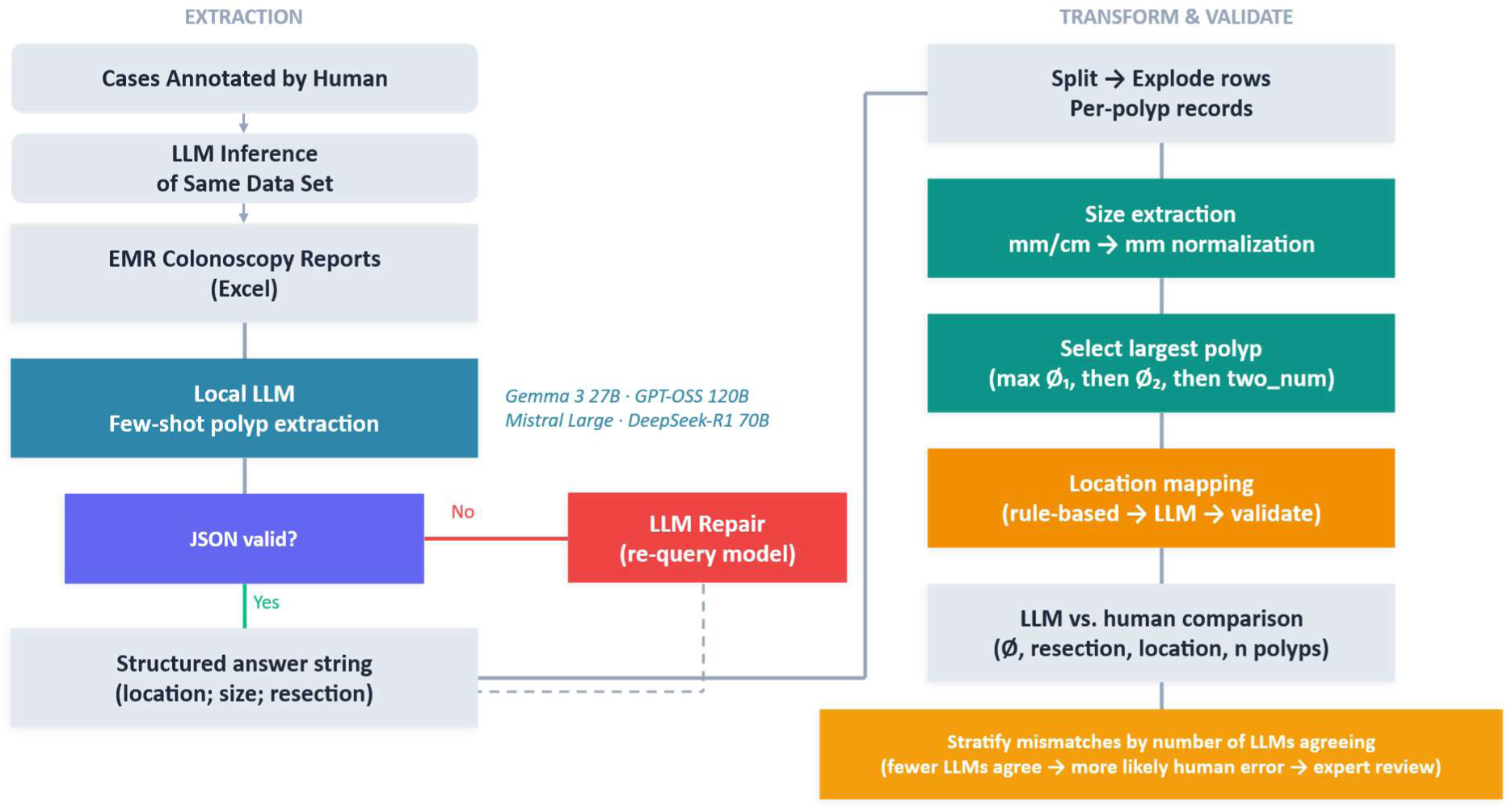
Detailed extraction and validation pipeline. The flowchart illustrates the two-stage processing pipeline used to generate structured polyp annotations from free-text colonoscopy reports. During the extraction (left), each EMR report was processed by four locally hosted LLMs (Gemma 3 27B, GPT-OSS 120B, Mistral Large, DeepSeek-R1 70B) using few-shot prompting. Model outputs were parsed as JSON; malformed responses were automatically re-queried for LLM-based repair, yielding a structured answer string containing location, size, and resection information. In the transformation and validation stage (right), multi-polyp reports were split into per-polyp records, size values were extracted and normalized to millimeters, and the largest polyp was selected based on a hierarchical criterion (maximum primary diameter, then secondary diameter). Anatomical locations were mapped through a combined rule-based and LLM-assisted classification step. The resulting LLM-derived annotations were then compared against the human reference labels across five categories (location, diameter 1, diameter 2, resection status and multiple polyps), and mismatches were stratified by the number of LLMs agreeing with the human annotator to prioritize cases for review.

**Supplementary Figure 3:**
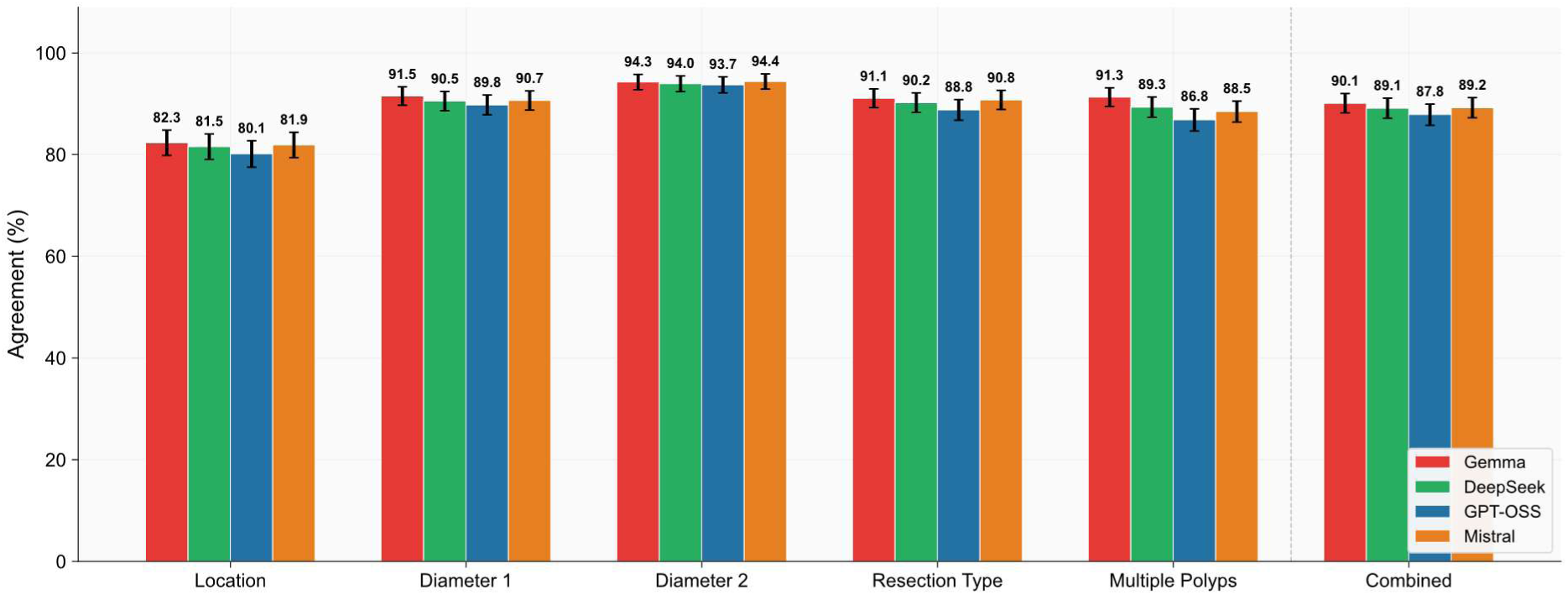
Agreement between each LLM and human annotations. Percentage agreement between each of the four LLMs (Gemma, DeepSeek, GPT-OSS, Mistral) and the human annotations across five extraction categories (en-bloc resection, multiple polyps, diameter 1, diameter 2, and anatomical location) as well as the combined agreement across all categories. Bar heights represent percentage agreement; error bars indicate 95% confidence intervals.

**Supplementary Figure 4:**
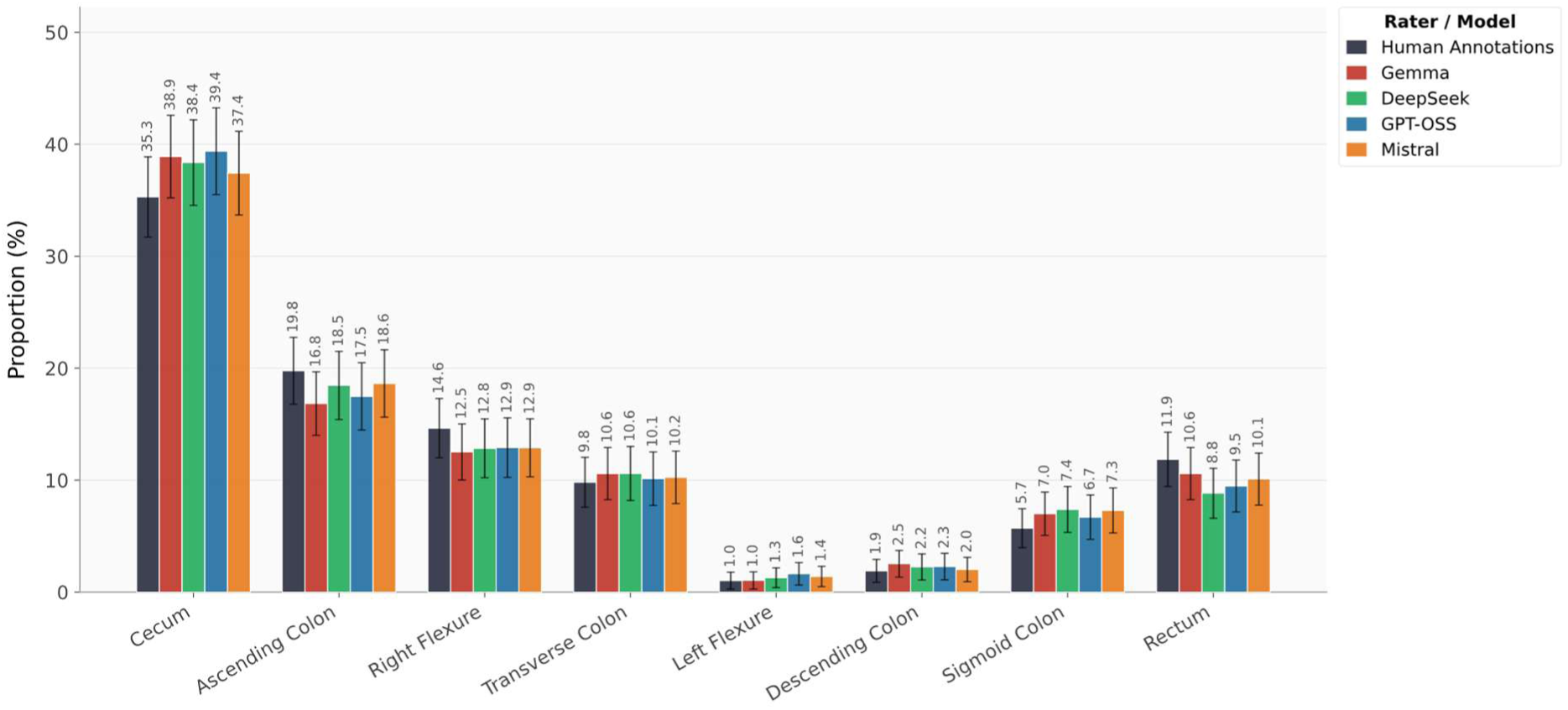
Distribution of polyp locations. Distribution of polyp locations across anatomical segments as annotated by the human annotator and four LLMs (Gemma, DeepSeek, GPT, Mistral). Bar heights represent the percentage of polyps assigned to each colonic location; error bars indicate 95% confidence intervals. All five raters produced broadly similar distribution patterns, with the cecum as the most frequently reported location (35.1–41.1%), followed by the ascending colon, right flexure, and rectum, while distal segments (transverse colon, left flexure, descending colon, sigmoid colon) each accounted for fewer than 10% of cases. The overall concordance in distributional shape across human and LLM annotations suggests that all raters captured the same underlying anatomical distribution, with only minor variation in the relative proportions assigned to individual segments.

**Supplementary Figure 5:**
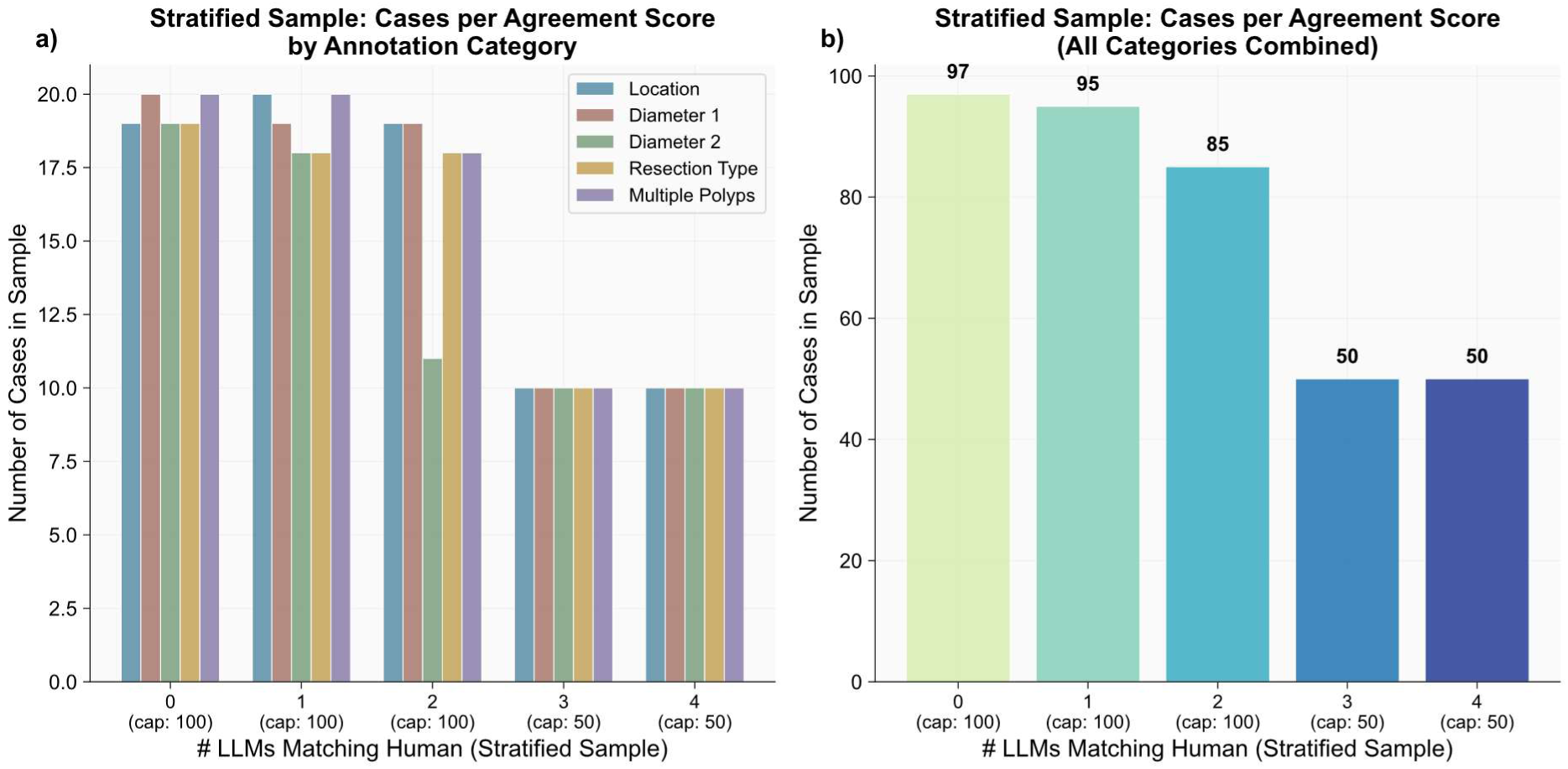
Stratified evaluation sample. The dataset is stratified for review by sampling up to 100 instances (20 per category) for agreement scores 0, 1, and 2, and 50 instances (10 per category) for agreement scores 3 and 4. a) shows the number of sampled cases per annotation category within each agreement group. b) displays the aggregated totals across all categories.

**Supplementary Figure 6:**
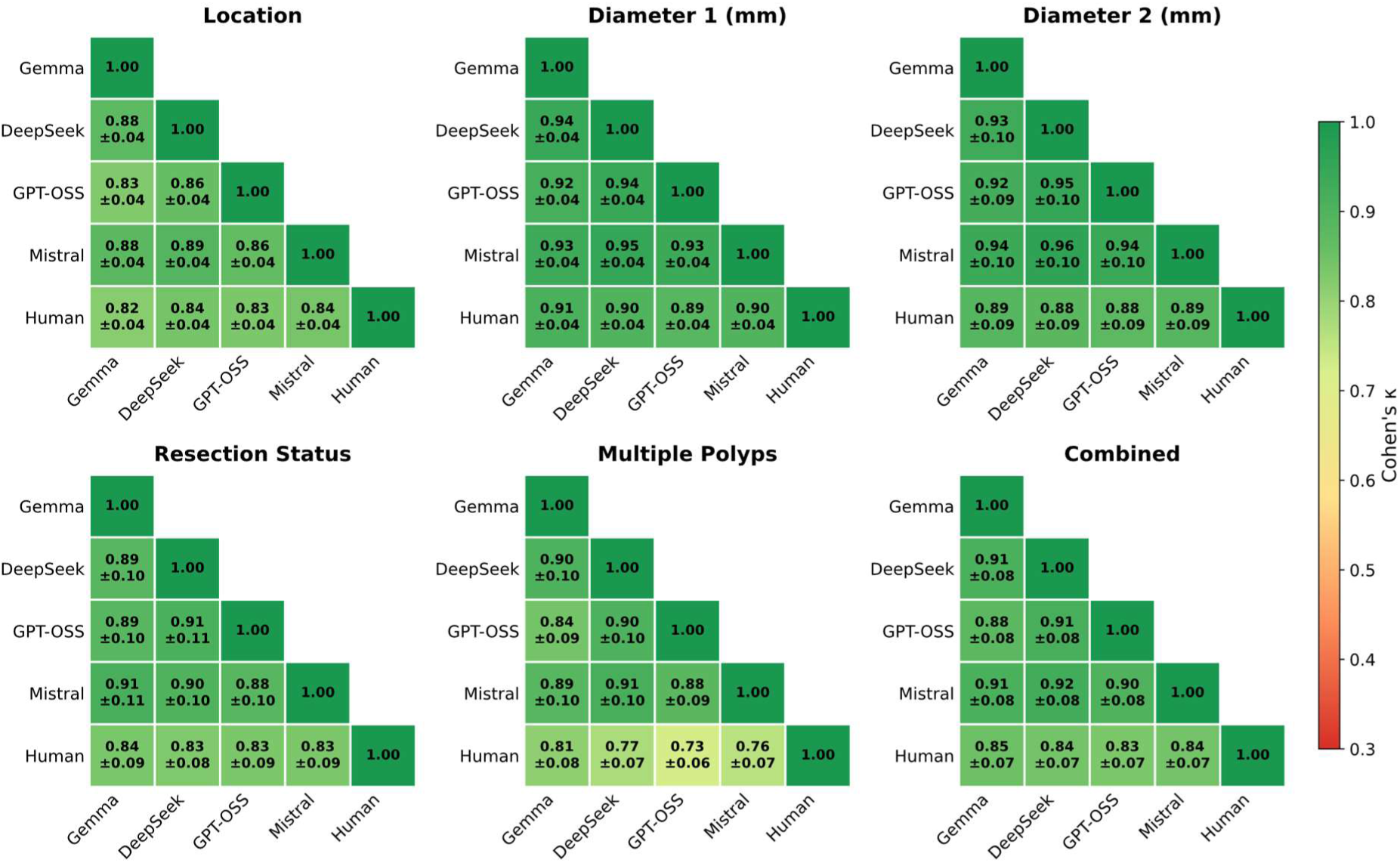
Pairwise Cohen’s κ heatmaps for all five raters. Pairwise Cohen’s κ heatmaps for Gemma, DeepSeek, GPT-OSS, Mistral, and the human annotator across the five extraction categories (location, diameter 1, diameter 2, resection status and multiple polyps) as well as the combined values are displayed. Values are reported as mean ± 95% confidence interval.

**Supplementary Figure 7:**
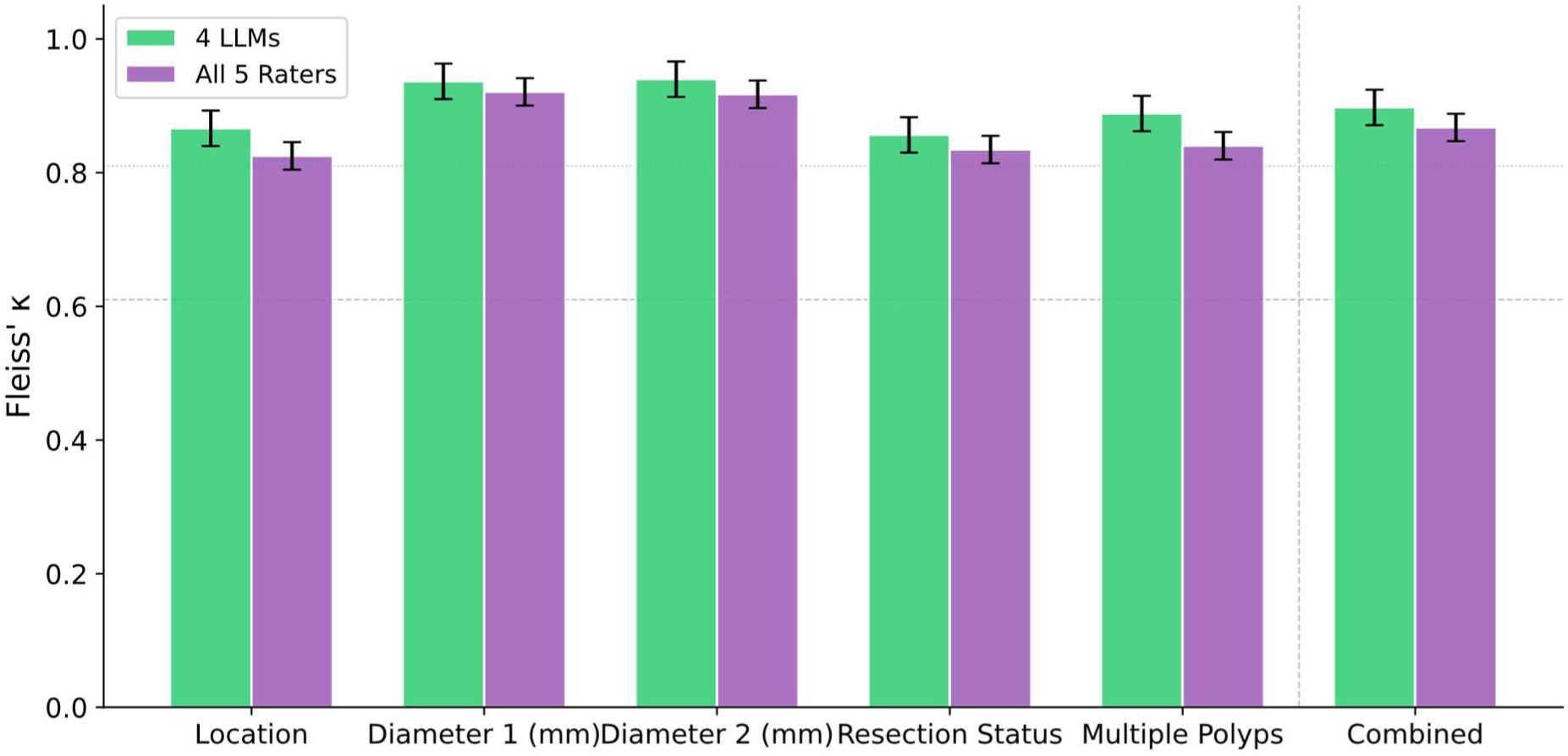
Fleiss’ κ by extraction category. Each category was computed for the four LLMs alone (green) and for all five raters including the human annotator (purple), with 95% confidence intervals. Excluding the human annotator increased overall group agreement only modestly, though pairwise comparisons revealed more substantial differences.

**Supplementary Figure 8:**
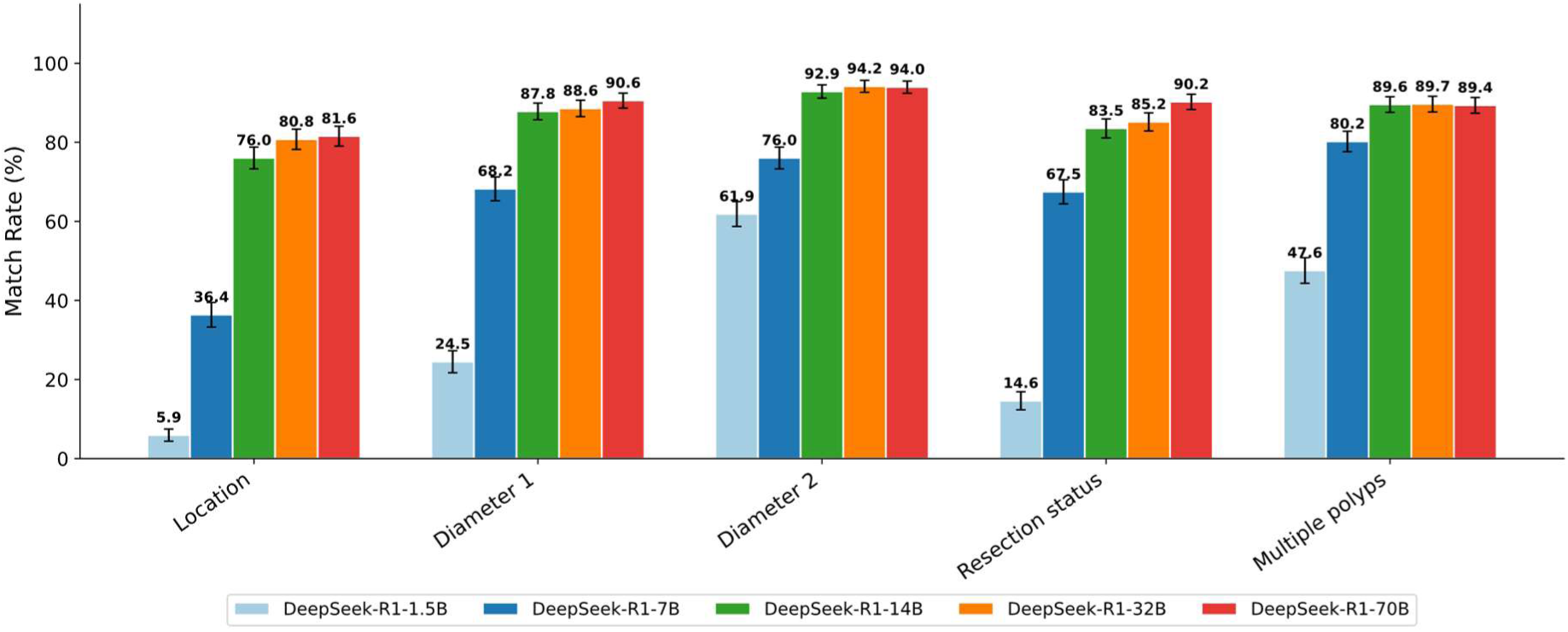
Match rates for different model sizes (DeepSeek-R1). Match rates (%) between LLM-extracted and human-annotated polyp characteristics across DeepSeek-R1 model sizes (1.5B, 7B, 14B, 32B, 70B) for five extraction targets: anatomical location, diameter 1, diameter 2, resection status, and multiple polyps. Match rates improve substantially with increasing model size, with the steepest gains observed between the 1.5B and 14B variants. Performance increases markedly from 1.5B to 7B and again from 7B to 14B, but improvements beyond 14B are only marginal. The 1.5B model shows notably poor performance across all categories (5.9–61.9%), with the 14B model approaching the performance ceiling for most tasks. The highest match rates were observed for diameter 2 and the lowest for location, consistent with the pattern observed for Gemma 3. Error bars represent 95% confidence intervals. Of note, the 70B variant is distilled into a Llama 3.3 70B base, whereas the smaller variants (1.5B–32B) use Qwen2.5 as their base architecture [16]; all variants are fine-tuned using reasoning data generated by the full DeepSeek-R1 671B model.

**Supplementary Figure 9:**
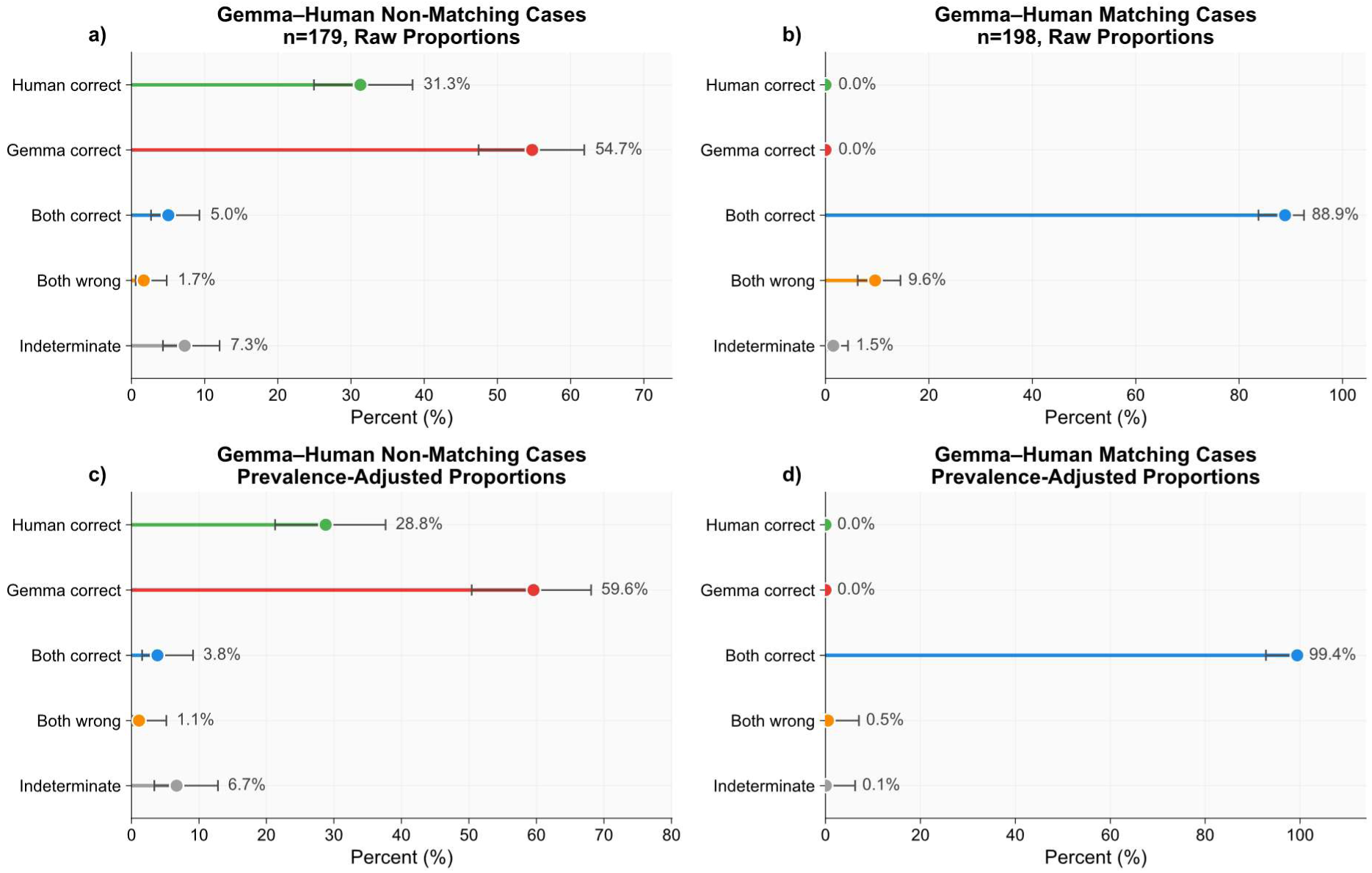
Second-reviewer adjudication results for Gemma annotations. Panels (a) and (b) show raw proportions; panels (c) and (d) show prevalence-adjusted proportions that correct for the stratified sampling design of the evaluation dataset. Left panels (a, c) display cases where Gemma and the human annotator disagreed (n=179); right panels (b, d) display cases where both agreed (n=198). Error bars represent 95% Wilson confidence intervals. Categories indicate whether the experienced reviewer judged the human annotator as correct, the LLM (Gemma) as correct, both as correct, both as wrong, or the case as indeterminate.

**Supplementary Figure 10:**
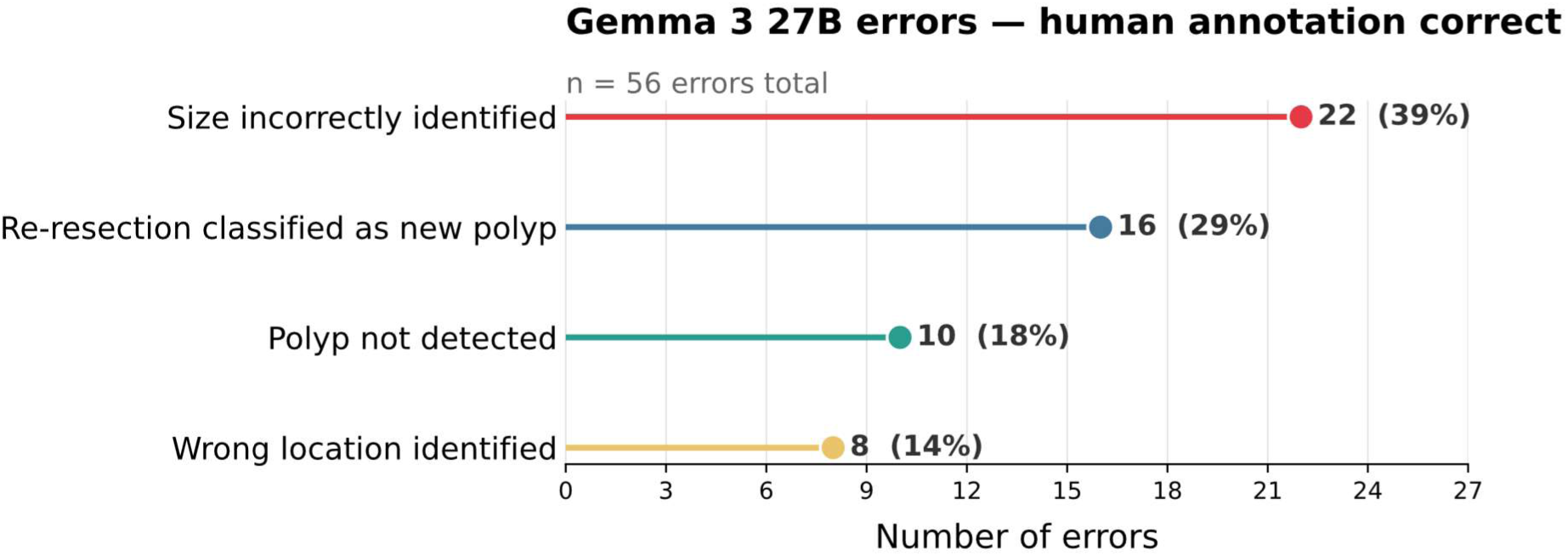
Qualitative analysis of Gemma 3 27B misclassifications where human annotation was correct (n = 56). Cases where Gemma 3 27B misclassified while the human annotator was correct were categorized by failure reason, determined through qualitative review of model output and corresponding endoscopy reports. Four error categories were identified: size incorrectly identified, re-resection classified as new polyp, polyp not detected, wrong location identified.

**Supplementary Table 1:** Variables extracted from unstructured data. Overview of the five clinical variables extracted from colonoscopy reports, including their data types and value ranges. Variables 1-4 were extracted for the largest polyp only.

| **Variable** | **Data type** | **Value** | **Description** |
| --- | --- | --- | --- |
| Anatomical location<br>(1) | Categorical | e.g., cecum,<br>ascending colon,<br>transverse colon, ... | Location of polyp in<br>the colon |
| Primary diameter<br>(2) | Continuous | Number in mm | Diameter of the<br>largest polyp.<br>Primary diameter is<br>the largest if two are<br>given |
| Secondary diameter<br>(3) | Continuous | Number in mm | Perpendicular<br>diameter (set to 0 if<br>not available) |
| Resection technique<br>(4) | Binary | En-bloc/piecemeal | Method used for<br>polyp removal |
| Multiple polyps<br>removed (5) | Binary | Yes / No | Whether more than<br>one polyp was<br>removed |

**Supplementary Table 2: Few-shot prompt**

English translation of few-shot prompt for extracting structured data from unstructured German language endoscopy reports. The same prompt was used in its original German form for all 4 LLMs.

“Few-Shot Prompt for extracting removed lesions

Task: Extract from an endoscopic report text only the adenomas that were actually removed. Output the results as a structured list, each line in the format: Location; Size; Method

Output only the results, no additional explanation or introduction! If an adenoma was only biopsied or left in place, do not include it in the list. If multiple adenomas were removed, list each as a separate line.

Assignment Rules

- Removed = phrasing such as removed as a whole (en-bloc) or in parts (piecemeal). If a polyp was removed piecemeal but overall completely, it counts as piecemeal. If no further details are given, it is en-bloc.

- Not removed = phrasing such as biopsied, left in place, left in place on withdrawal, only biopsied → omit these adenomas.

- Size: if stated in the text, use exactly as written; otherwise mark as not specified.

- Location: as precise as possible (e.g., sigmoid, ascending colon, transverse colon, rectum, left flexure, right flexure, cecum, 18 cm from the anal verge).

report: unremarkable. In the sigmoid a polyp bud, biopsy II. Smooth advancement to the cecum. Here, behind the valve, a 60 × 90 mm adenoid tumor is found. Randomization to STEP in group 3. Good lifting on submucosal injection with HydroJet. Piecemeal complete resection, no bleeding. Small remnants treated with APC until no tumor is macroscopically visible. At the edge of the resection surface a slit-shaped muscularis defect develops, closed with 4 clips, aspiration of free air. Retrieval in net (I)

Expected output:

- Cecum; 60 × 90 mm; piecemeal

report: Advancement to the unremarkable end of the Hartmann’s stump at 28 cm from the anal verge. At 18 cm from the anal verge the approx. 1 × 1 cm adenoma, which after submucosal injection with HAES/TB is removed in toto. Through the stoma advancement to the cecal pole, where a small adenoma at the cecal pole is removed with forceps (biopsy I). A further approx. 1.5 × 1 cm flat adenoma opposite the IC valve is biopsied and left in place (biopsy II). Directly on the IC valve a approx. 3 × 2 cm suspicious adenoma is only biopsied (biopsy III). In the ascending colon, 3 sessile adenomas of approx. 5 mm, 8 mm, and 10 mm are found in close proximity, which after submucosal injection with HAES/TB are removed in toto with a snare (biopsy IV). The adenomas are retrieved in a net. A further adenoma in the transverse colon measuring just over 1 cm is left in place on withdrawal. The remaining colon appears unremarkable.

Expected output:

- 18 cm from the anal verge; 1 × 1 cm; en-bloc

- Cecal pole; not specified; en-bloc

- Ascending colon; 5 mm; en-bloc

- Ascending colon; 8 mm; en-bloc

- Ascending colon; 10 mm; en-bloc

report:

Unremarkable on retroflexion. With stiffening advancement to the cecal pole and intubation of the terminal ileum over 15 cm. Unremarkable ileal mucosa. Two folds distal to the IC valve, two 1 cm flat polyps are found which after submucosal injection can be completely removed with the snare; only one can be retrieved (biopsy III). In the distal ascending colon two 1 cm polyps are found, retrieved together after snare resection (biopsy IV). Left flexure removal of one further polyp 1 cm snare (biopsy V) and in the descending colon a pedunculated polyp removed with snare (biopsy VI). Overall pronounced diverticulosis with maximum in the sigmoid.

Expected output:

- IC valve; 1 cm; en-bloc

- IC valve; 1 cm; en-bloc

- Ascending colon; 1 cm; en-bloc

- Ascending colon; 1 cm; en-bloc

- Left flexure; 1 cm; en-bloc

- Descending colon; not specified; en-bloc

report:

Between 5 and 8 cm from the anal verge a roughly 35 mm flat elevated tumor, alongside 3 satellite lesions each approximately 7 mm, of which one is removed and retrieved (III), one removed, and one left in place. The main lesion is injected and circumcised with HybridKnife, bleeding during the procedure managed with Coag-Grasper. Patient becomes restless, therefore procedure abbreviated and resection in 3 portions with snare. Hemostasis with 3 clips, coagulation. Smooth advancement to the cecum. Throughout the colon, but particularly frequently in the sigmoid, numerous large non-irritated diverticula. In the sigmoid at approximately 25 cm patchy redness (biopsy II). In the descending colon (110 cm on insertion) a flat elevated adenoid tumor 12 mm is injected and resected, alongside a further 5 mm directly resected (I).

Expected output:

- 5–8 cm from the anal verge; 35 mm; piecemeal

- 5–8 cm from the anal verge; 7 mm; en-bloc

- 5–8 cm from the anal verge; 7 mm; en-bloc

- Descending colon; 12 mm; en-bloc

- Descending colon; 5 mm; en-bloc

report:

Unremarkable sigmoid with numerous diverticula and very angulated. Otherwise smooth passage to the cecum. From the IC valve 2 × biopsy (I) to rule out adenoma. Alongside a flat elevated adenoid tumor 25 × 35 mm, randomization to STEP, good lifting on submucosal injection, subtotal resection in 1 piece (II), remainder treated with APC.

Expected output:

- IC valve; 25 × 35 mm; piecemeal

Return your answer exclusively as a valid JSON object. Exactly two fields are allowed:

1. “reasoning”: a brief external summary of your reasoning (no internal thoughts, no <think>)
2. “answer”: a single string containing the final answer. No arrays, no objects, plain text only.

Formatting rules for “answer”:

- Each removed lesion on its own line

- Format of each line: “<Location>; <Size>; <Removal method>”

- Lines separated by “\n”

- No additional fields, no extra structures.

Important:

- Output absolutely no text outside the JSON.

- No explanations, no comments, no internal thoughts, no <think>.

- Only a single JSON object.”

**Supplementary Table 3: Location normalization prompt**

The locations were mapped to standardized format using the following prompt if simple matching algorithm failed.

“You are a medical assistant for endoscopic documentation. Assign the following location description to one of the predefined categories.

Location description: **Insert_Location_Here**

Rules:

- IC valve, cecal pole, appendix → Cecum
- Right or left flexure, transverse colon → corresponding flexure or transverse colon
- Linea dentata, anal canal, ampulla, anorectal → Rectum
- Rectosigmoid junction → Sigmoid
- Ignore prefixes such as distal, proximal
- If no unambiguous assignment is possible → return the original description unchanged Valid categories:

**VALID_LOCATION_STR**

Return a JSON object with two fields:

- “location”: exactly one of the categories listed above (exact spelling)
- “reason”: brief justification Return only valid JSON, nothing else.”

**Valid locations are the following:**

VALID_LOCATIONS_STR = [

“cecum-left flexure”, “indeterminate”, “right flexure”,

“left flexure”, “see_distance”, “cecum”, “ascending colon”, “transverse colon”, “descending colon”, “sigmoid colon”,

“rectum”,

]

